# The efficacy and safety of the prasugrel, ticagrelor, and clopidogrel dual antiplatelet therapies following an acute coronary syndrome: A systematic review and Bayesian network meta-analysis

**DOI:** 10.1101/2023.08.12.23294021

**Authors:** Stephen A. Kutcher, Leah Flatman, Rachelle Haber, Nandini Dendukuri, Sonny Dandona, James M. Brophy

## Abstract

**Background:** The dual-antiplatelet therapies (DAPT) of clopidogrel, prasugrel, or ticagrelor in concomitant use with acetylsalicylic acid are the contemporary treatment regimens for acute coronary syndromes (ACS). Systematic comparative effectiveness and safety analyses currently lack clinically meaningful interpretations of the summarized evidence.

**Methods:** We systematically searched MEDLINE, EMBASE, CENTRAL, and clinicaltrials.gov for randomized controlled trials (RCTs) that reported on either the efficacy or safety between clopidogrel, prasugrel, or ticagrelor DAPTs in ACS patients. The primary efficacy endpoint was a composite of all-cause mortality, a recurrent non-fatal myocardial infarction, or non-fatal stroke. The primary safety endpoint was study-reported major bleeding events. A Bayesian network meta-analysis was performed using a generalized linear model logit transformation with a log-transformation of ‘time’ for varying lengths of study follow-up. Studies published in either English or French with a minimum of 6 months of follow-up and a “low” rating from the Cochrane risk of bias assessment tool were included in the main analyses. Fixed and random effects models fit was assessed by the deviance information criterion (DIC) and node-splitting methods were used to assess the consistency of direct and indirect network evidence. An HR >0.9 and <1.11 were set as our clinically important thresholds, and represented the range of practical equivalence (ROPE).

**Results:** From a total of 15,232 articles identified, 138 were selected for full-text review. From a total of 29 identified RCT’s, 17 trials, representing 57,814 subjects, were identified as a “low” risk of bias and were included in the final Bayesian network meta-analysis. Compared to clopidogrel, prasugrel and ticagrelor reduced major acute coronary events (MACE) endpoints by a median of 13% (Hazard ratio [HR]PC, 0.87; 95% credible interval [95% CrI]: 0.74, 1.06) and 5% (HRTC, 0.95; 95% CrI: 0.81, 1.14), respectively. The HR posterior distributions estimated that prasugrel had a 67.5% chance of producing a clinically meaningful – greater than 10% (HR<0.9) – decrease in the risk of MACE outcomes, while ticagrelor only had a 22.4% chance of exceeding the clinically important threshold. The primary safety outcome found prasugrel (HRPC, 1.23; 95% CrI: 1.04, 1.40) and ticagrelor (HRTC, 1.07; 95% CrI: 0.99, 1.17) DAPTs to be associated with a median increase in events relative to clopidogrel. This translates to a probability of a clinically meaningful increase (HR>1.11) in major bleeding of 83.7% for prasugrel and 67.7% for ticagrelor, when compared to clopidogrel.

**Conclusion:** When compared with ACS patients assigned to clopidogrel, prasugrel and ticagrelor were associated with moderate and modest probabilities respectively in clinically meaningful MACE reductions. Prasugrel and ticagrelor had high and modest probabilities respectively of clinically meaningful increases in bleeding. Despite guideline recommendations, the net clinical benefit for these drugs compared to clopidogrel appears uncertain.

## INTRODUCTION

Dual antiplatelet therapy (DAPT), consisting of the concomitant use of acetylsalicylic acid (ASA) and clopidogrel, is standard secondary prevention following an acute coronary syndrome (ACS) hospitalization. The most recent American,^1^ Canadian,^2^ and European^3^ ACS guidelines suggest the use of ticagrelor and prasugrel DAPT over clopidogrel for the treatment of ACS, unless patients are at high-risk of bleeding. These ACS guidelines largely rely on evidence from two large multinational randomized controlled trials, PLATO^4^ and TRITON-TIMI-38,^5^ that reported a clinical efficacy benefit of ticagrelor (hazard ration[HR]: 0.84, 95% confidence interval [CI]: 0.77 to 0.92; P<0.001) and prasugrel (HR: 0.81, 95%CI: 0.73 to 0.90; P<0.001), respectively, when compared to clopidogrel.

However, if one desires to personalize DAPT choice, for example by considering the geographic region where the patient is treated the situation becomes less clear. Two publications report on the pre-specified PLATO regional analysis. One study includes the 1,413 patients from the United States (HRUS: 1.27, 95%CI: 0.92 to 1.75)^6^ and another by the Food and Drug Administration (FDA) on the whole 1,814 North American subjects (HRNA: 1.25, 95%CI: 0.93 to 1.67),^7^ find no benefit for the efficacy outcome, major acute coronary events (MACE), from ticagrelor over clopidogrel. In fact, the sub-analyses suggest the potential for an increase in MACE outcomes with reported HRs greater than one. These North American sub-population estimates deviate substantially from the overall pooled effect reported for the entire study population (HRPLATO: 0.84).

It may be argued that there is *one* overall “true” treatment effect (a fixed effect) and that the results within North American patients are simply due to random chance. Alternatively, it can be viewed that the observed HR discrepancies are due to inherent and meaningful population differences and healthcare practices across the study regions; patients were recruited from a total of 43 and 21 countries in PLATO and TRITON-TIMI-38, respectively. It is plausible that the effect estimates from these large RCTs rather come from a distribution of effects (a random effect) and the added uncertainty from regional variability should be accounted for in the analyses.^8^ A recent discussion^9^ describing the utility of Bayesian analytical methodologies, re-analyzed the PLATO data applying a hierarchical (random effects) approach accounting for the regional differences. They reported a pooled HR of 0.87 with a 95% credible interval (CrI) from 0.70 to 1.15. The superiority of ticagrelor over clopidogrel became less confident using the hierarchical model, as the interpretation of the effect estimate was not robust – the 95%CrI crosses the null (HR=1.0) – to a simple change in a modelling assumption (fixed versus random effects). This signal was not dismissed in the FDA report,^7^ as multiple reviewers made calls for further evidence prior to the drug’s approval.

The purpose of this paper is to systematically search the literature for RCTs comparing clopidogrel, ticagrelor, or prasugrel in ACS patients that include MACE and/or major bleeding outcomes with a minimum of 6 months follow-up. A Bayesian network meta-analysis will be performed to compare the direct and indirect evidence on the efficacy and safety across the three DAPTs, with a particular focus on identifying RCT evidence from North American study populations.

## METHODS

Following the PRISMA extension for network meta-analyses^10^ we performed a systematic search of EMBASE, MEDLINE, the Cochrane controlled register of trials (CENTRAL), and clinicaltrials.gov, last updated on August 1st, 2022. The PRISMA checklist can be found in the appendix. The complete search strategy is also described in the appendix but briefly, we searched for RCTs using the terms “acute coronary syndrome” or “myocardial ischemia” or “unstable angina” or “ST segment elevation myocardial infarction” or “non-ST segment elevation myocardial infarction” or “unstable angina” and related keywords. Additionally, we searched for “dual antiplatelet therapy” or “DAPT” or a combination of “prasugrel” and “clopidogrel” and “ticagrelor”. A hand-search of the references from relevant articles was further performed. Article inclusion criteria were: (1) patients diagnosed with ACS with or without percutaneous coronary intervention (PCI) or coronary artery bypass graft (CABG) surgeries; (2) patients assigned to two of the following DAPT treatment regimens: clopidogrel, ticagrelor, or prasugrel – with a minimum of 6-month planned treatment duration and follow-up; (3) randomized controlled trials of human subjects; (4) reporting at on the main composite outcome of MACE (defined as mortality, recurrent MI, or stroke); and, (5) studies published in either English or French.

Due to likely variations in reporting definitions of major acute coronary events (MACE), we adopted a “hard” MACE definition for the primary efficacy outcome, a composite measure of reported all-cause mortality, non-fatal MI, or stroke. Secondary efficacy endpoints include the individual MACE outcomes, all-cause mortality, non-fatal MI, or non-fatal stroke, as well as cardiovascular-caused mortality, if available. The primary safety endpoint of interest was major bleeding as defined by the trial. Our expectation was that most studies will report Thrombolysis In Myocardial Infarction (TIMI) major bleeding, PLATO-defined major bleeding, and Bleeding Academic Research Consortium (BARC) types 3-5, but we will accept the bleeding definition provided by the included study (Table 1). The secondary safety endpoint included minor bleeding as defined by TIMI, PLATO, and BARC (types 1 and 2), or other reported minor bleeding definitions.

**Table 1:**
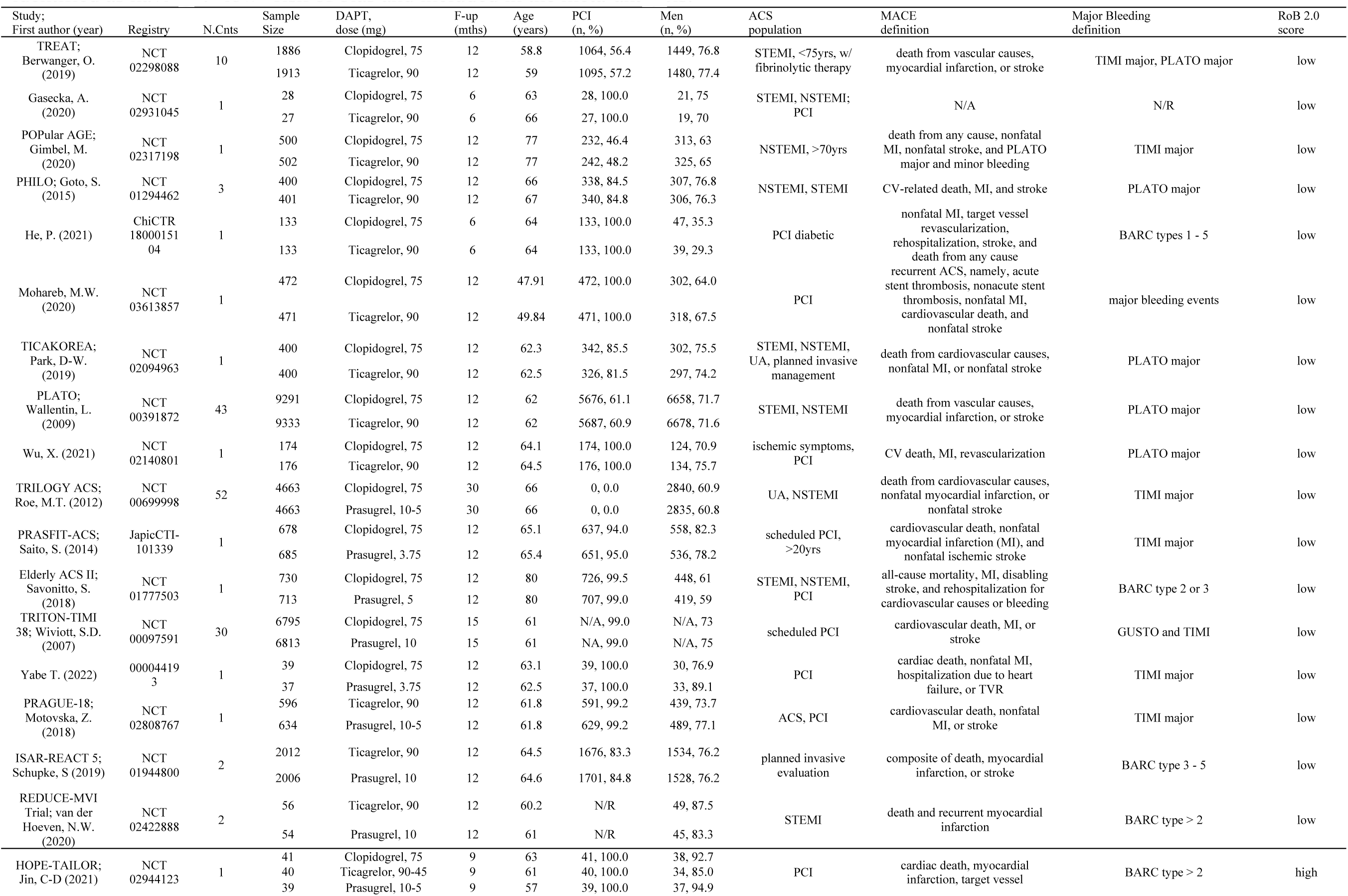
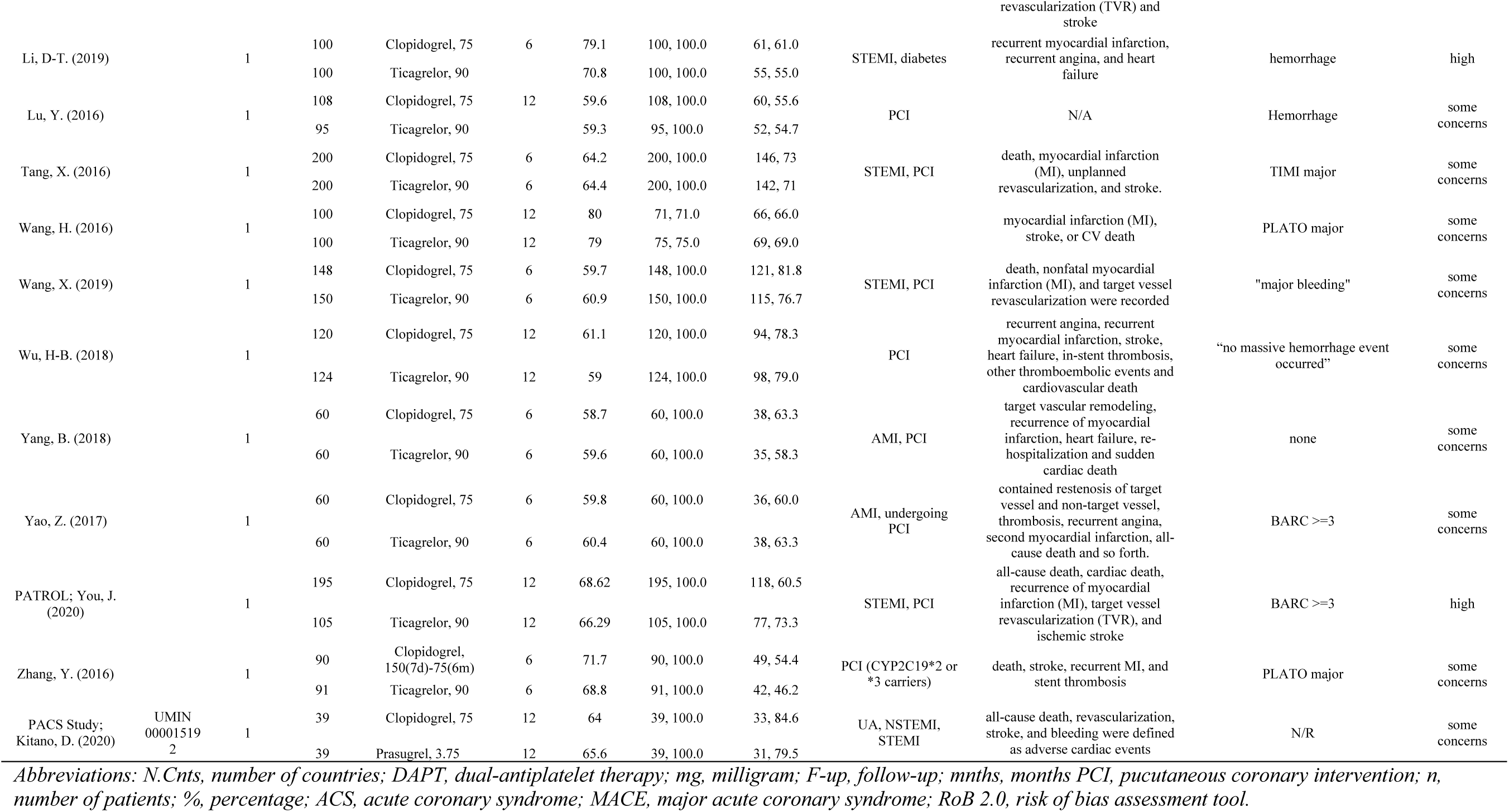
The descriptive characteristics of the twenty-nine studies identified from the systematic review. The first seventeen studies were identified as having a “low” risk of bias and included in the meta-analysis.

Data extraction was performed by S.K. and quality assessment of eligible studies was carried out simultaneously by S.K., L.F., and R.H. Information on the year of publication, sample size, the loading and maintenance dosages of treatment regimens, length of planned treatment duration, the mean age, body mass index (BMI), proportion of males, and those who underwent PCI or CABG, as well as the description of the study population and country of origin were collected. The Cochrane revised tool to assess risk of bias in randomized trials (RoB 2.0)^11^ was used to assess allocation concealment, blinding of participants and trial personnel, reporting on completeness and selection of outcomes, and other biases. The scoring system for each component of the RoB 2.0 includes a “low”, “some concerns” or “high” risk of bias. Studies with a score of “low” across all five categories were considered low risk of bias and included in the analyses. A funnel plot will be used to visually assess publication bias.

## STATISTICAL ANALYSIS

Mixed treatment, or network meta-analyses (NMA), is a technique that allows for the synthesis of multiple treatment strategies from independent research studies, using direct (head-to-head) and indirect (common node/comparator) comparisons.^12,13^ Direct effects come from the head-to-head comparisons of the treatments. Indirect effects come from comparing the non-head-to-head comparisons by using the information from across the studies that have a common node, or comparator.

Due to the potential for variations in reporting practices of the outcomes of interest, we chose a Bayesian random effect generalized linear model with a logit outcome transformation.^14^ The further potential for trials reporting differing lengths of follow-up, a complementary log-log (cloglog) link function was included to the model, by adding a log(follow-up time) variable, to account for these variations in follow-up times. The cloglog model estimates log-hazard ratios and assumes that hazards are constant over the duration of follow-up and is homogenous across each trial.^14^

Both fixed and random effects binomial likelihood models with a cloglog link function were applied to the data using the “*gemtc*” R package^15^ with Bayesian posterior distribution sampling from the Markov Chain Monte Carlo (MCMC) which was estimated via the Just Another Gibbs Sampler (JAGS) using the “*rjags*” R package.^16^ The models were estimated using a minimum of 50,000 iterations with a 10,000 burn-in period run on 3 chains. Model diagnostics were verified by examining traceplots (via monitoring of MCMC sampling), posterior distributions and a multivariate potential scale reduction factor (mpsrf) above 1.1.^17,18^ In the case of model inconsistencies, the burn-in and number of iterations were increased. The *gemtc* default non-informative priors were used for all model parameters. Statistical analyses were performed using R version 4.0.2 from the R Project for Statistical Computing.^19^

The random effects (RE) model was selected as our primary analysis as the fixed effect (FE) assumption of a homogeneous population-level effect across multiple study populations seems improbable. Nonetheless, the assessment of model fit was performed using the lowest deviance information criterion (DIC) – the sum of the models’ residual deviance and its’ leverage – which penalizes models with more parameters. Some have suggested that when a FE and a RE model are within five DIC units,^14^ the model with fewer parameters (FE) should be favoured. To be comprehensive, we also reported the FE estimates for comparative purposes. Further, the network consistency – the agreement between the direct, indirect and total network evidence across the pair-wise comparisons – was visually assessed using the node splitting methods.^12^

The use of a Bayesian analytical approach allows for direct probability statements for the interpretation of the summarized evidence for the efficacy and safety endpoints. It eliminates the need for null hypothesis significance testing and the commonly misunderstood P-value.^20^ Instead, probability statements, about the likely benefit or harm of the antiplatelet strategy relative to the reference, clopidogrel, will be presented. In addition, probability statements will be provided for a net “clinical” benefit or harm, which has been arbitrarily set at a 10% decrease (HR=0.9) or an equivalent increase (HR=1.11) in the effect estimate. The proportion of the posterior distribution that falls within the region of practical equivalence (ROPE), between HR = [0.9, 1.11], will also be reported.^21^ In short, we are presenting readers the proportion of the HR posterior distribution that lies above, below, and between the “important” clinical thresholds.

As the study material consisted only of previously published material IRB and/or ethics committee approvals were not required.

## RESULTS

A total of 9,196 titles and abstracts were screened following the removal of duplicates and research abstracts from the 15,232 records identified from the systematic search. This resulted in a full-text assessment of 138 articles from which 29 trials,^4,5,22–48^ for a total of 60,278 study participants, met the inclusion criteria for the primary efficacy or safety endpoint. A flowchart (Figure 1) presents a summary of the screening process, and a description of the included studies are presented in Table 1. Seventeen^4,5,22–36^ of the 29 articles were assessed a “low” risk of bias score, across all five RoB 2.0 assessment categories, and included in the Bayesian network meta-analysis.

**Figure 1:**
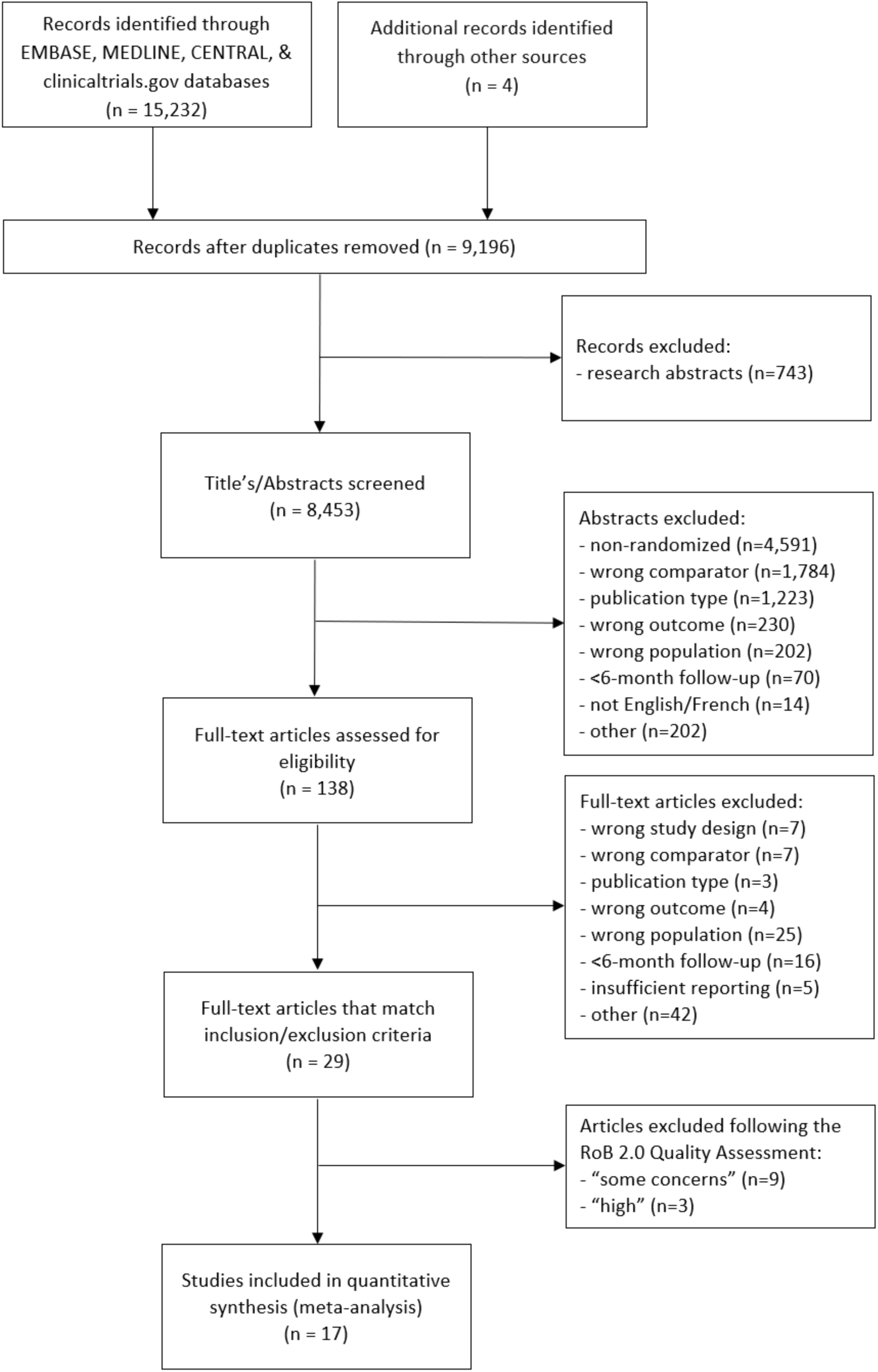
A flowchart describing the systematic screening results.

For this triangular NMA a total of nine (52.9%),^4,22–29^ of the included RCTs compared the DAPTs’ clopidogrel with ticagrelor, while five (29.4%)^5,30–33^ trials compared clopidogrel to prasugrel and three (17.6%)^34–36^ contrasted ticagrelor with prasugrel (Figure 2). The majority of the included RCTs built-in 12 months of follow-up time, with two^23,26^ studies following patients for 6 months, and another two trials^5,30^ following subjects for up to 15 and 30 months, respectively. The average or median age of the included study populations ranged from 47.9 years ^27^ to 80 years,^32^ in 13 (76.5%) of the median age was between 60 to 69. Twelve (70.6%) of the study populations had planned invasive ACS management and reported that greater than 80% of patients underwent a PCI. All but one RCT (5.9%)^26^ reported studying a majority (>60%) of male patients.

**Figure 2:**
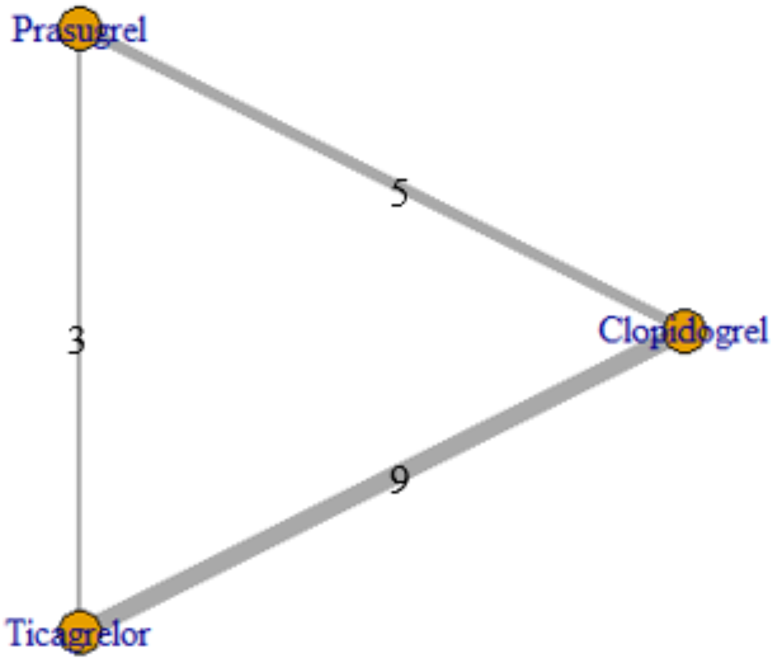
The network structure of the Bayesian meta-analysis for the 17 trials identified from the systematic review.

### Primary outcomes

#### Major acute coronary events

The seventeen included studies, with a total of 57,814 subjects, reported a total of 6,897 (11.9%) “hard” MACE outcomes. The RE model (DICRE=56.60) was considered a better overall fit relative to the FE model (DICFE=65.29). Visual inspection of the node-splitting models favoured the RE model, with more concordance between the direct, indirect, and network evidence (Figure 3E-F), suggesting heterogeneity of the efficacy estimates across the included pairwise RCTs.

**Figure 3:**
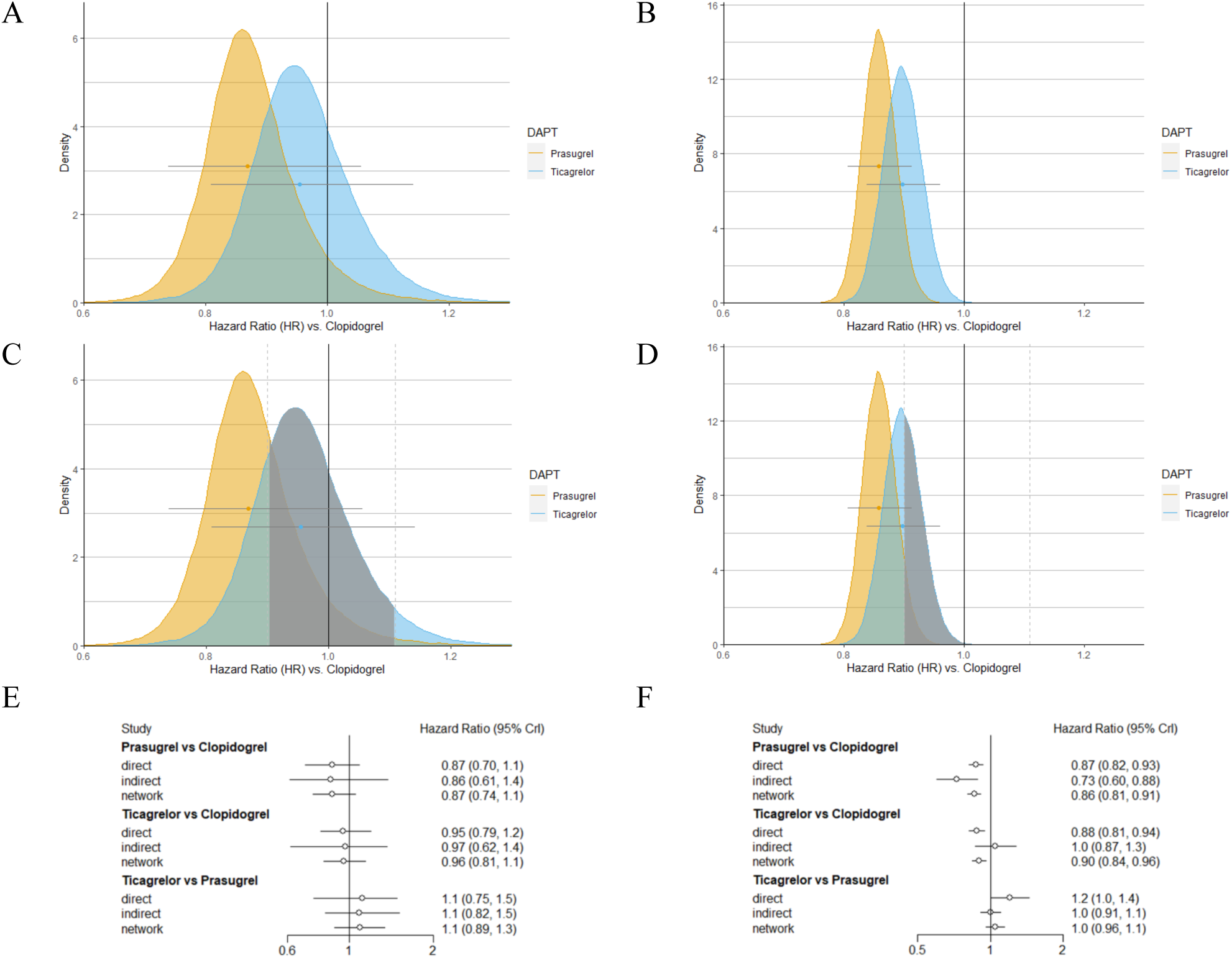
The posterior distributions for major acute coronary events (MACE) comparing clopidogrel (reference) to ticagrelor (blue) and prasugrel (yellow) using a [**A**] random effects (RE) and [**B**] fixed effects models. The range of practical equivalence (ROPE) is highlighted in grey across the RE [**C**] and FE [**D**] models. The node splitting results for MACE in the RE [**E**] and FE [**F**] models. *Abbreviations: DAPT, dual anti-platelet therapy; CrI, credible interval*.

The ticagrelor to clopidogrel HR estimand for MACE outcomes was 0.95 (95% CrI: 0.81, 1.14) (Table 2; Figure 5). This translated to 22.4% of the posterior distribution being below the 10%, clinically important reduction threshold (HR<0.9) (Table 2; Figure 3C). The majority of the HRTC distribution (73.2%) for MACE outcomes fell within the ROPE (HR = [0.9 to 1.11]).

**Table 2:**
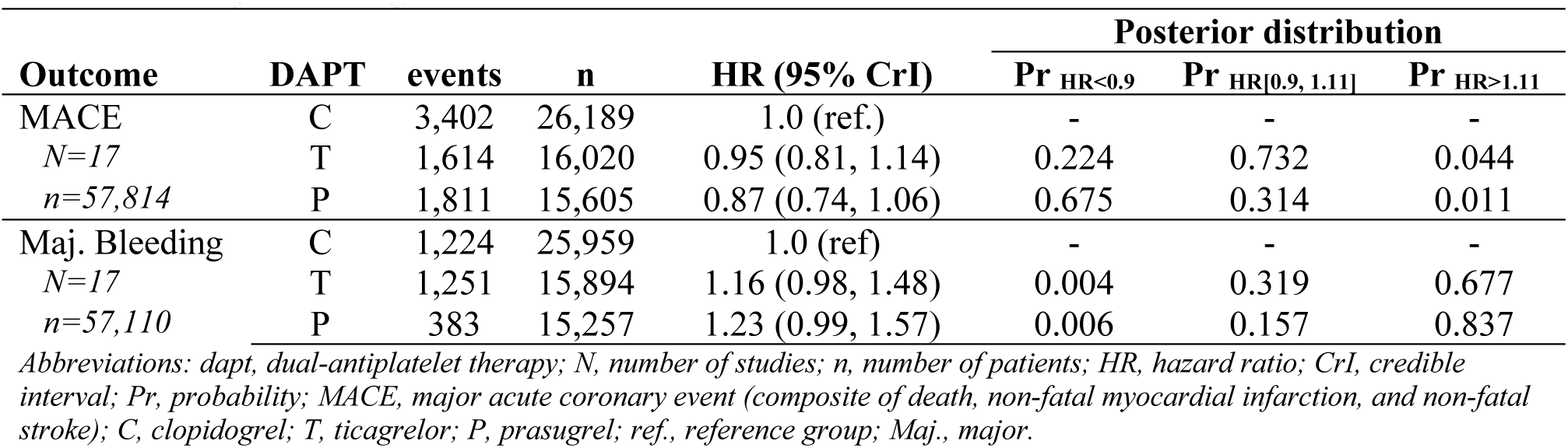
The results of the random effects Bayesian network meta-analyses for the primary outcomes of MACE and major bleeding.

| Outcome | DAPT | events | n | HR (95% CrI) | Posterior distribution |  |  |
| --- | --- | --- | --- | --- | --- | --- | --- |
|  |  |  |  |  | Pr<sub>HR<0.9</sub> | Pr<sub>HR[0.9, 1.11]</sub> | Pr<sub>HR>1.11</sub> |
| MACE | C | 3,402 | 26,189 | 1.0 (ref.) | - | - | - |
| *N=17* | T | 1,614 | 16,020 | 0.95 (0.81, 1.14) | 0.224 | 0.732 | 0.044 |
| *n=57,814* | P | 1,811 | 15,605 | 0.87 (0.74, 1.06) | 0.675 | 0.314 | 0.011 |
| Maj. Bleeding | C | 1,224 | 25,959 | 1.0 (ref.) | - | - | - |
| *N=17* | T | 1,251 | 15,894 | 1.16 (0.98, 1.48) | 0.004 | 0.319 | 0.677 |
| *n=57,110* | P | 383 | 15,257 | 1.23 (0.99, 1.57) | 0.006 | 0.157 | 0.837 |

Abbreviations: *dapt*, dual-antiplatelet therapy; *N*, number of studies; *n*, number of patients; *HR*, hazard ratio; *CrI*, credible interval; *Pr*, probability; *MACE*, major acute coronary event (composite of death, non-fatal myocardial infarction, and non-fatal stroke); *C*, clopidogrel; *T*, ticagrelor; *P*, prasugrel; *ref.*, reference group; *Maj.*, major.

The prasugrel to clopidogrel HR estimand for MACE outcomes was, 0.87 (95% CrI: 0.74, 1.06) (Table 2; Figure 5). This extrapolated to 67.5% of the posterior distribution of the HR falling beyond the meaningful clinical threshold (HR<0.9) for a reduction in MACE outcomes, with 31.4% of the distribution captured within the ROPE (Table 2;Figure 3C).

#### Major bleeding

The primary safety outcome, reported major bleeding (Table 1), was available in the 17 (57,110 patients) trials for a total of 2,858 (5.0%) recorded events. The RE model (DICRE=52.82) was considered a similar fit overall relative to the FE model (DICFE=54.40). Visual inspection of the node-splitting models found superior concordance between estimates from the direct, indirect, and network in the RE model (Figure 4 E-F), across all 3 pairwise comparisons.

**Figure 4:**
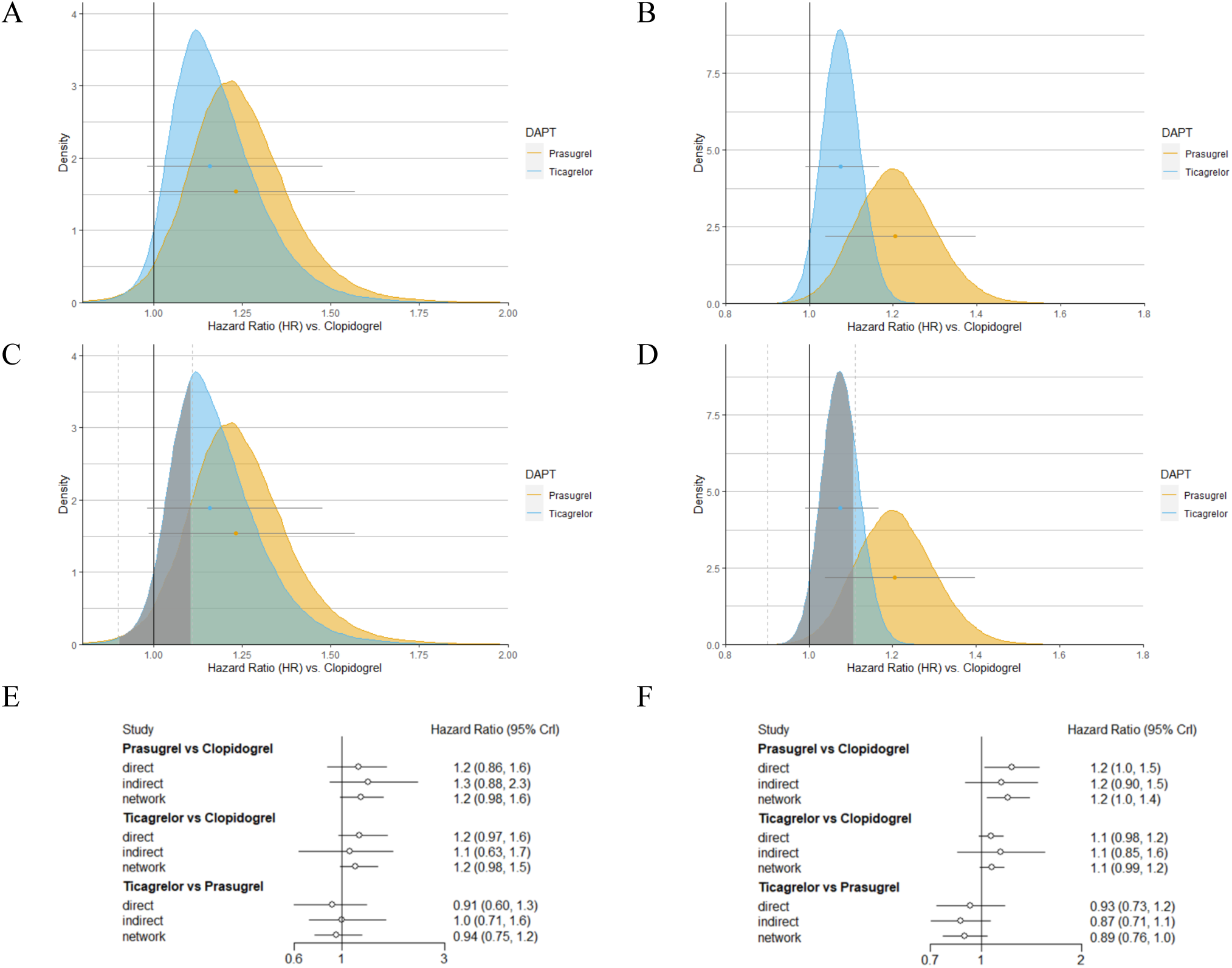
The posterior distributions for major bleeding events comparing clopidogrel (reference) to ticagrelor (blue) and prasugrel (yellow) using a [**A**] random effects (RE) and [**B**] fixed effects models. The range of practical equivalence (ROPE) is highlighted in grey across the RE [**C**] and FE [**D**] models. The node splitting results for major bleeding in the RE [**E**] and FE [**F**] models. *Abbreviations: DAPT, dual anti-platelet therapy; CrI, credible interval*.

In comparison to clopidogrel, the ticagrelor HR estimand for major bleeding, as defined within the included trial, was 1.16 (95% CrI: 0.98, 1.48) (Table 2; Figure 5). This translated to 67.7% of the posterior surpassing a clinically meaningful increase in major bleeding events in those exposed to clopidogrel, relative to ticagrelor, while 31.9% of the posterior fell within the ROPE (Table 2; Figure 4E).

**Figure 5:**
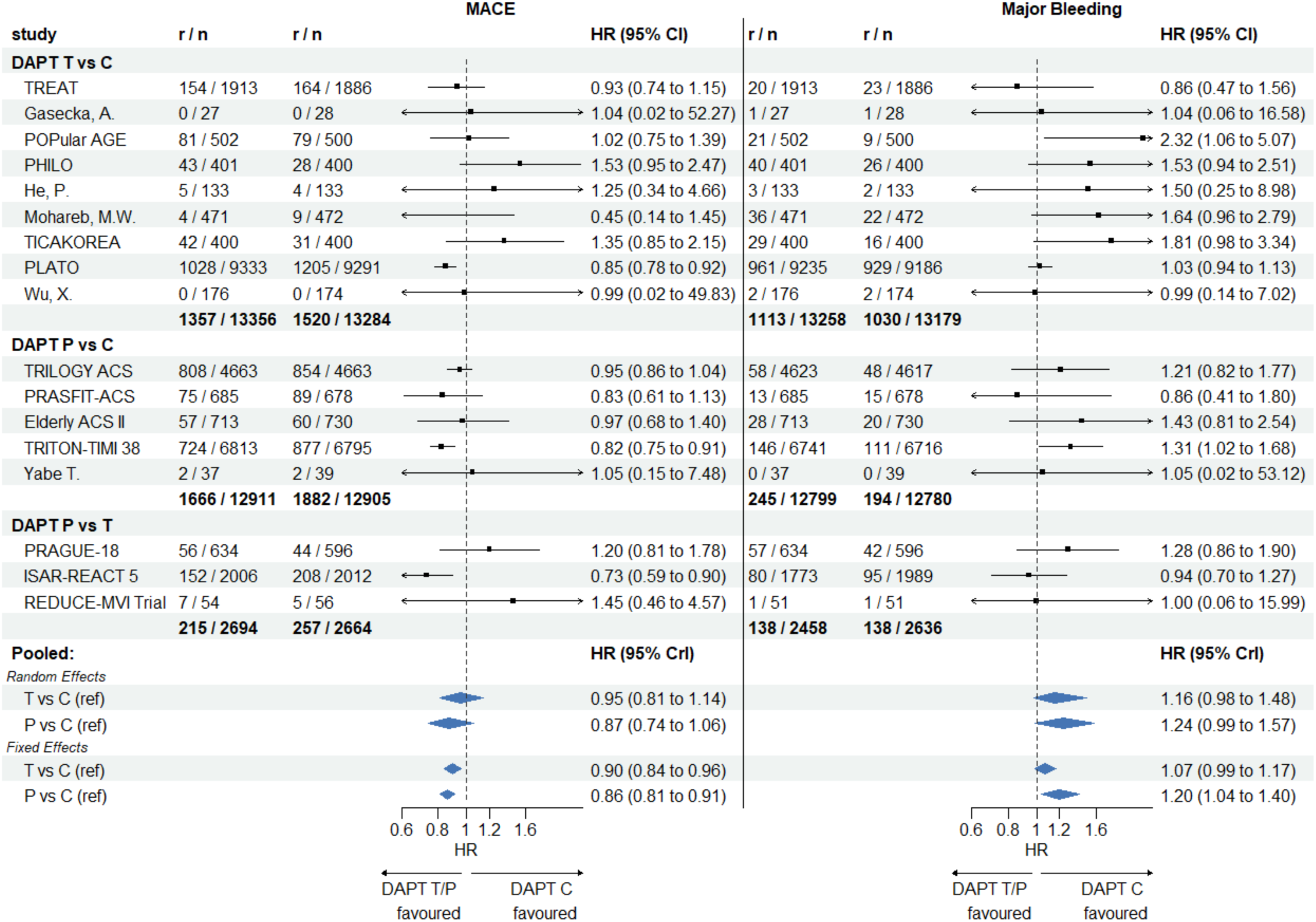
Forest plot of summary of the Bayesian network meta-analyses for MACE and major bleeding events in patients taking clopidogrel compared to ticagrelor and clopidogrel. *Abbreviations: MACE, major acute coronary events; r, number of events; n, number of subjects per study arm; HR, hazard ratio; CI, confidence interval; CrI, Bayesian credible interval; DAPT, dual-antiplatelet therapy; C, clopidogrel; T, ticagrelor; P, prasugrel; ref, reference group*.

The prasugrel to clopidogrel HR estimand for major bleeding events was 1.23 (95% CrI: 0.99, 1.57) (Table 2; Figure 5). This extrapolated to 83.7% of the posterior surpassing the clinically important threshold for an increase in bleeding, with 15.7% found within the ROPE (Table 2; Figure 4E).

### Secondary outcomes

#### All-cause mortality

The 17 RCTs reported a total of 2,688 (4.6%) mortality outcomes. The FE (DIC=51.38) and RE (DIC=51.60) models had comparable fits, though, node-splitting models identified some potential inconsistency (disagreement) between the indirect, direct, and network model estimates. The HR estimand for all-cause mortality, comparing ticagrelor to clopidogrel, was 0.87 (95% CrI: 0.75, 1.07), where 64.2% of the posterior fell below the clinically important reduction threshold (HR<0.9). Meanwhile, the HR estimand, for prasugrel relative to clopidogrel, was 0.94 (95% CrI: 0.79, 1.14), with 30.5% of the posterior found below the clinically meaningful threshold (Supplemental Table 2).

#### Cardiovascular-related mortality

Fifteen trials (57,438 patients) reported 2,182 (3.80%) cardiovascular-related deaths (CV-deaths). In comparison to clopidogrel, the HR estimand of CV-related mortality for ticagrelor was 0.85 (95% CrI: 0.73, 1.04). It was estimated that 75.3% of the posterior was below the clinically meaningful threshold. Meanwhile, the HR estimand comparing prasugrel to clopidogrel was 0.90 (95% CrI: 0.76, 1.07), where 51.6% of the posterior distribution was below the threshold of clinical importance (Supplemental Table 2).

#### Myocardial infarction

The 17 RCTs (57,148 patients) reported a total of 3,539 (6.1%) non-fatal MIs. The RE model (DIC=52.09) was borderline a statistical improvement (<5 DIC units), relative to the FE model (DIC=57.00). However, moderate qualitative inconsistency from the node-splitting method reinforced the RE model assumptions. When compared to clopidogrel, the HR for MI outcomes for ticagrelor was found to be 0.94 (95% CrI: 0.76, 1.17), with 32.8% of the posterior being represented below a clinically meaningful threshold and 61.2% within the ROPE. The HR estimand, comparing prasugrel to clopidogrel, was estimated as 0.81 (95% CrI: 0.65, 1.00) (Supplemental Table 2). A total of 87.4% of the prasugrel posterior was beyond the threshold of a clinical important reduction in MI’s.

#### Stroke

A total of 12 studies (56,521 patients) reported on 670 (1.2%) stroke outcomes. Node-splitting models suggested the network to be unstable with inconsistent direct, indirect, and network estimates across all three (C-T, C-P, and T-P) pair-wise comparisons. The posterior distributions for ticagrelor (HR, 1.02; 95% CrI: 0.72, 1.33) and prasugrel (HR, 0.88; 95% CrI: 0.64, 1.18), when compared with clopidogrel are presented (Supplemental Table 2).

#### Minor bleeding

Twelve (49,677 patients) RCTs reported a total of 1,352 (2.7%) minor bleeding events. Node-splitting models, under both FE and RE assumptions, suggested the network to be somewhat unstable regarding the inconsistent direct, indirect, and network estimates across all three (C-T, C-P, and T-P) pair-wise comparisons. The RE posterior distributions for minor bleeding outcomes for ticagrelor (HR, 1.35; 95% CrI: 1.08, 1.71) and prasugrel (HR, 1.42; 95% CrI: 1.08, 1.94), when compared with clopidogrel are provided (Supplemental Table 2).

The FE results for both the primary and secondary, efficacy and safety outcomes were reported in the appendix (Supplemental Tables 1 & 3).

## DISCUSSION

The main efficacy findings from this large (n=57,814) Bayesian network meta-analysis of DAPT strategies following an ACS hospitalization are: (1) ticagrelor is associated with a small decrease in MACE outcomes but that is unlikely to provide a meaningfully clinically important (>10%) reduction in endpoints; (2) prasugrel, when compared with clopidogrel, is associated with a larger (13%) reduction in MACE but only a moderate probability (67.5%) of providing a clinically relevant reduction. The primary safety findings from the NMA include: (3) that ticagrelor compared to clopidogrel is associated with an increase in major bleeding events, but only a small probability (21.3%) that this exceeds a clinically meaningful increase. And, (4) prasugrel, when compared with clopidogrel, is also associated increased in major bleeding with a moderate to high probability (85.3%) that this exceeds a clinically meaningful threshold.

The results from our Bayesian NMA do not fully align with the recommendations of the published North American guidelines^1,2^ and other previous NMAs.^49,50^ While the summary estimates for prasugrel and ticagrelor show a reduction in MACE outcomes, they do not demonstrate the same degree of certainty as the PLATO and TRITION-TIMI trials or NMAs, on which the guidelines rely. Using classic confidence limits of 95%, neither prasugrel (95%CrI: 0.74, 1.06) nor ticagrelor (95%CrI: 0.81, 1.14) establish superiority over clopidogrel. Further, our results estimate that there is a 32.5% and 77.6% chance that prasugrel and ticagrelor, respectively, do not provide a clinically meaningful reduction in MACE endpoints when compared with clopidogrel. There is some concordance in the guidelines with respect to the primary safety outcome, major bleeding, and exercising caution when using these more potent platelet inhibitors – prasugrel (95% CrI: 0.99, 1.17) and ticagrelor (95% CrI: 1.04, 1.40) – in subjects with a higher risk of bleeding. It remains a balance of weighing the benefits and harms of the treatments and secondary patient outcomes.

The strength of our findings are built upon the mixed-treatment models of the BNMA, which optimizes evidence synthesis, under certain assumptions, by permitting the use of both direct and indirect evidence across the three guidelines approved DAPTs – clopidogrel, ticagrelor, and prasugrel –in ACS patients. The inclusion of only low-risk RCTs bolsters the internal validity of the current findings, which is supported by a large ACS patient population (n=57,814). The Bayesian approach in this study allows for direct probability statements to be made for the interpretation of the treatment effect estimates, thus avoiding the often-misinterpreted P-value. This BNMA goes beyond others^49,50^ by also providing probability statements regarding the chances of observing a clinically meaningful change (>10%) in the benefits or harms, an important factor for clinical decision-makers.

As with all research, our study comes with limitations. Our findings *do not* account for the potential challenges previously identified internal modelling decisions (RE vs FE) found in the large multinational RCTs^4,5,22,30^ included in this BNMA. This study assumed that the within study variance was properly specified in the individual RCTs when the between-study and between-study-arm variance were modelled. Meaning, we likely underestimated the size of the credible intervals of the posterior estimates. Further, our BNMA models included only those studies that received a “low” risk of bias score across the five RoB 2.0 bias assessment tool. Twelve studies (2,464 patients)^37–40,42–48^ received at minimum a “some concerns” reviewer score in at least one RoB 2.0 assessment category. Of note, all twelve excluded studies recruited 200 or fewer patients, and only two^37,48^ of the excluded RCTs (16.7%) reported a RCT registration number versus all seventeen (100%) of the included trials. Conditioning the analyses on could also impact funnel plot symmetry for bias assessment. Although our analyses did not elucidate any concerns regarding the distribution of effect sizes (Supplemental Figure 1), it has been suggested that this is difficult to assess with fewer than 10 studies^51^ – T vs. C (n=9); P vs. C (n=5); and, P vs. T (n=3). Transitivity (between network arms) and homogeneity (between study arms) can be assessed qualitatively by comparing the similarity of study populations and treatments. There were some minor differences across some of the study populations, such as age, percent PCI, percent male characteristics, and outcome definitions, which could represent some between study heterogeneity of which could impact the transitivity and consistency assumptions. Overall, the populations represented patients with acute cardiac symptoms (STEMI, NSTEMI, and/or UA) and received well defined medications. Reporting differences for the efficacy outcome was minimized through adopting a “hard” MACE outcome, to include the more objective clinical outcomes of death, MI, or stroke.

In conclusion, ticagrelor was estimated to have a 22.4% chance of decreasing MACE and a probability of 21.3% of increased major bleeding, while prasugrel was associated with a 67.5% probability in reducing MACE and an 85.3% increased chance of major bleeding outcomes, when compared with ACS patients assigned to clopidogrel. The results of this BNMA do not provide robust evidence regarding the superiority of ticagrelor and prasugrel, over clopidogrel, as the 95% credible intervals include the null and are substantially captured within the ROPE region. We were also unable to identify a recent RCT on North American subjects. Further research is required to better understand the heterogeneity in the effects of DAPTs within diverse ACS populations, especially with limited North American evidence.

## Data Availability

All data produced in the present study are available upon reasonable request to the authors

## APPENDIX

**Supplemental Table 1:**
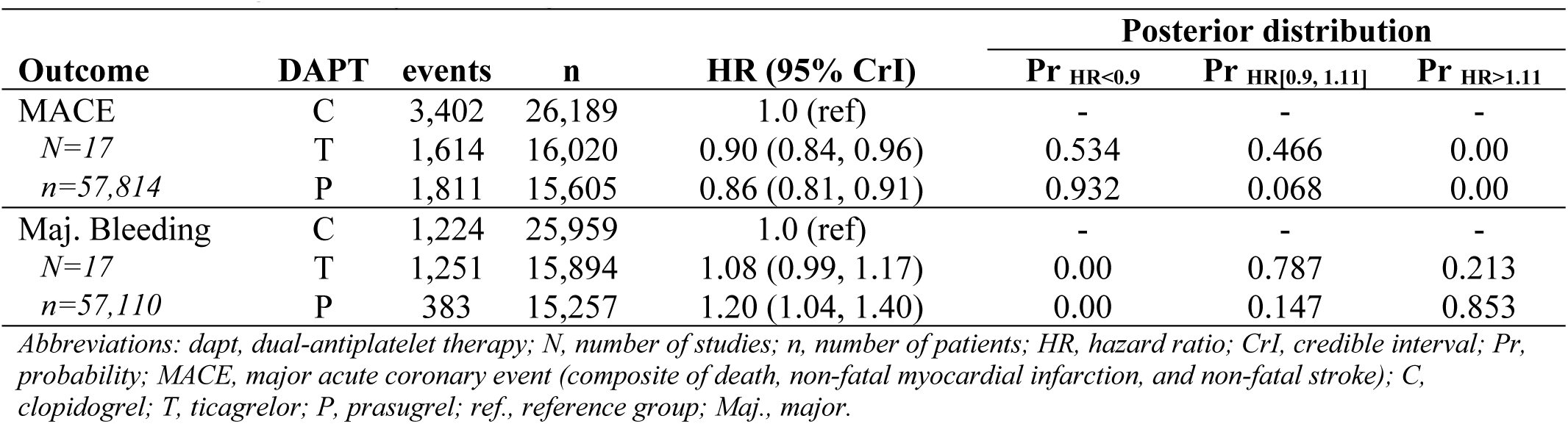
The results of the fixed effects Bayesian network meta-analyses for the primary outcomes of MACE and major bleeding.

**Supplemental Table 2:** The random effect results of secondary outcomes from the Bayesian network meta-analyses.

| Outcome | DAPT | events | n | HR (95% CrI) | Posterior distribution |  |  |
| --- | --- | --- | --- | --- | --- | --- | --- |
|  |  |  |  |  | Pr<sub>HR<0.9</sub> | Pr<sub>HR[0.9, 1.11]</sub> | Pr<sub>HR>1.11</sub> |
| Mortality | C | 1,299 | 26,189 | 1.0 (ref) | - | - | - |
| *N=17* | T | 661 | 16,020 | 0.87 (0.75, 1.07) | 0.642 | 0.344 | 0.014 |
| *n=57,814* | P | 728 | 15,605 | 0.94 (0.79, 1.14) | 0.305 | 0.658 | 0.037 |
| CV Mortality | C | 1,076 | 26,056 | 1.0 (ref) | - | - | - |
| *N=15* | T | 547 | 15,831 | 0.85 (0.73, 1.04) | 0.753 | 0.239 | 0.008 |
| *n=57,438* | P | 559 | 15,551 | 0.90 (0.76, 1.07) | 0.516 | 0.473 | 0.011 |
| MI | C | 1,798 | 26,189 | 1.0 (ref) | - | - | - |
| *N=17* | T | 750 | 16,020 | 0.94 (0.76, 1.17) | 0.328 | 0.612 | 0.060 |
| *n=57,814* | P | 991 | 15,605 | 0.81 (0.65, 1.00) | 0.874 | 0.121 | 0.005 |
| Stroke | C | 305 | 25,543 | 1.0 (ref) | - | - | - |
| *N=15* | T | 203 | 15,373 | 1.02 (0.72, 1.33) | 0.199 | 0.552 | 0.249 |
| *n=56,521* | P | 162 | 15,605 | 0.88 (0.64, 1.18) | 0.551 | 0.388 | 0.061 |
| Min. Bleeding | C | 548 | 24,729 | 1.0 (ref) | - | - | - |
| *N=12* | T | 550 | 12,811 | 1.35 (1.08, 1.71) | 0.002 | 0.034 | 0.964 |
| *n=49,677* | P | 254 | 12,137 | 1.42 (1.08, 1.94) | 0.003 | 0.031 | 0.966 |

Abbreviations: dapt, dual-antiplatelet therapy; N, number of studies; n, number of patients; HR, hazard ratio; CrI, credible interval; Pr, probability; CV, cardiovascular; MI, non-fatal myocardial infarction, and non-fatal stroke); C, clopidogrel; T, ticagrelor; P, prasugrel; ref., reference group; Min., minor.

**Supplemental Table 3:** The fixed effect results of secondary outcomes from the Bayesian network meta-analyses.

| Outcome | DAPT | events | n | HR (95% CrI) | Posterior distribution |  |  |
| --- | --- | --- | --- | --- | --- | --- | --- |
|  |  |  |  |  | Pr<sub>HR<0.9</sub> | Pr<sub>HR[0.9, 1.11]</sub> | Pr<sub>HR>1.11</sub> |
| Mortality | C | 1,299 | 26,189 | 1.0 (ref) | - | - | - |
| *N=17* | T | 661 | 16,020 | 0.84 (0.76, 0.94) | 0.890 | 0.110 | 0.00 |
| *n=57,814* | P | 728 | 15,605 | 0.93 (0.83, 1.03) | 0.305 | 0.695 | 0.00 |
| CV Mortality | C | 1,076 | 26,056 | 1.0 (ref) | - | - | - |
| *N=15* | T | 547 | 15,831 | 0.83 (0.74, 0.93) | 0.918 | 0.082 | 0.00 |
| *n=57,438* | P | 559 | 15,551 | 0.90 (0.80, 1.01) | 0.517 | 0.483 | 0.00 |
| MI | C | 1,798 | 26,189 | 1.0 (ref) | - | - | - |
| *N=17* | T | 750 | 16,020 | 0.90 (0.81, 0.99) | 0.531 | 0.469 | 0.00 |
| *n=57,814* | P | 991 | 15,605 | 0.81 (0.74, 0.88) | 0.992 | 0.008 | 0.00 |
| Stroke | C | 305 | 25,543 | 1.0 (ref) | - | - | - |
| *N=15* | T | 203 | 15,373 | 1.06 (0.86, 1.29) | 0.059 | 0.632 | 0.309 |
| *n=56,521* | P | 162 | 15,605 | 0.91 (0.73, 1.12) | 0.471 | 0.498 | 0.031 |
| Min. Bleeding | C | 548 | 24,729 | 1.0 (ref) | - | - | - |
| *N=12* | T | 550 | 12,811 | 1.34 (1.18, 1.52) | 0.00 | 0.002 | 0.998 |
| *n=49,677* | P | 254 | 12,137 | 1.39 (1.15, 1.68) | 0.00 | 0.009 | 0.991 |

Abbreviations: dapt, dual-antiplatelet therapy; N, number of studies; n, number of patients; HR, hazard ratio; CrI, credible interval; Pr, probability; CV, cardiovascular; MI, non-fatal myocardial infarction, and non-fatal stroke); C, clopidogrel; T, ticagrelor; P, prasugrel; ref., reference group; Min., minor.

**Supplemental Figure 1:**
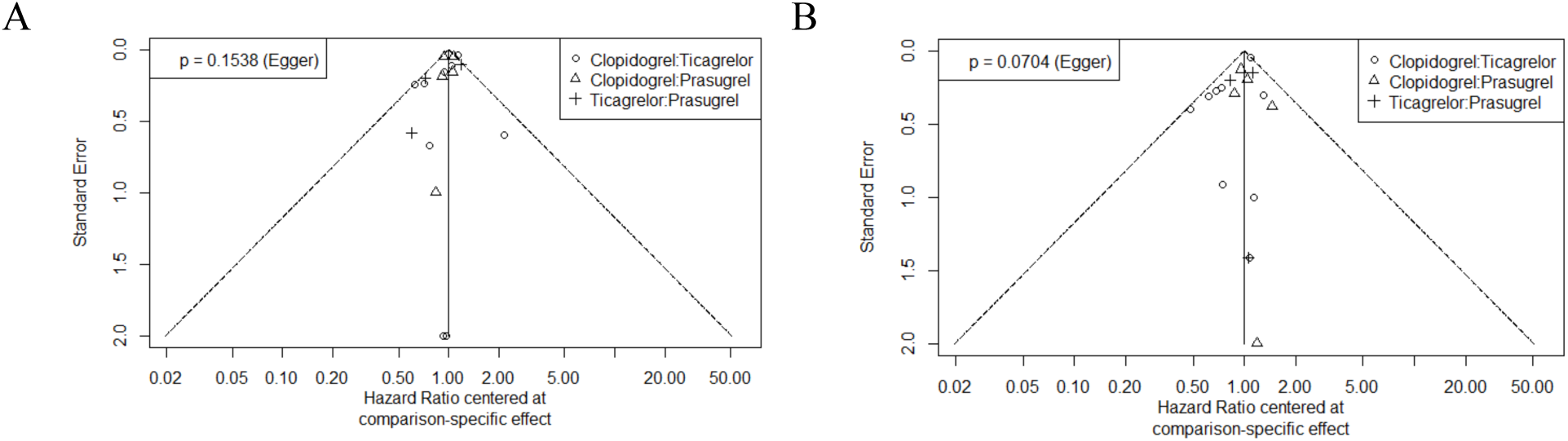
Publication bias funnel plots for major acute coronary events and major bleeding outcomes

### Search Strategies

#### Medline

1. ((randomized controlled trial or controlled clinical trial).pt. or randomized.ab. or randomised.ab. or placebo.ab. or drug therapy.fs. or randomly.ab. or trial.ab. or groups.ab.) not (exp animals/ not humans.sh.)
2. acute coronary syndrome*.mp. or Acute Coronary Syndrome/ or ((myocardial or heart) adj infarction*).mp. or acute mi.mp. or exp Myocardial Infarction/ or ((myocardial or heart muscle) adj isch?emi*).mp. or Myocardial Ischemia/ or unstable angina*.mp. or Angina, Unstable/ or STEMI.mp. or NSTEMI.mp.
3. (((percutaneous coronary or heart muscle) adj (intervention* or revasculari#ation*)) or pci).mp. or Percutaneous Coronary Intervention/
4. stent*.mp. or Stents/
5. coronary artery bypass*.mp. or Coronary Artery Bypass/
6. 2 or 3 or 4 or 5
7. (clopidogrel and ticagrelor).mp.
8. (clopidogrel and prasugrel).mp.
9. (prasugrel and ticagrelor).mp.
10. (dual antiplatelet* or dual anti platelet* or DAPT).mp.
11. 7 or 8 or 9 or 10
12. 1 and 6 and 11

#### Embase

1. crossover-procedure/ or double-blind procedure/ or randomized controlled trial/ or single-blind procedure/ or (random* or factorial* or crossover* or cross over* or placebo* or (doubl* adj blind*) or (singl* adj blind*) or assign* or allocat* or volunteer*).tw.
2. acute coronary syndrome*.mp. or acute coronary syndrome/
3. ((myocardial or heart) adj infarction*).mp. or heart infarction/
4. acute mi.mp.
5. myocardial isch?emi*.mp. or heart muscle ischemia/
6. unstable angina*.mp. or unstable angina pectoris/
7. non ST segment elevation myocardial infarction/
8. ST segment elevation myocardial infarction/
9. percutaneous coronary intervention.mp. or percutaneous coronary intervention/
10. heart muscle revascularization/ or ((percutaneous coronary or heart muscle) adj revasculari#ation*).mp.
11. percutaneous coronary revasculari#ation*.mp.
12. stent*.mp. or stent/
13. coronary artery bypass*.mp. or coronary artery bypass graft/
14. 2 or 3 or 4 or 5 or 6 or 7 or 8 or 9 or 10 or 11 or 12 or 13
15. (clopidogrel and prasugrel).mp.
16. (clopidogrel and ticagrelor).mp.
17. (prasugrel and ticagrelor).mp.
18. (dual antiplatelet* or dual anti platelet* or DAPT).mp.
19. 15 or 16 or 17 or 18
20. 1 and 14 and 19

#### CENTRAL

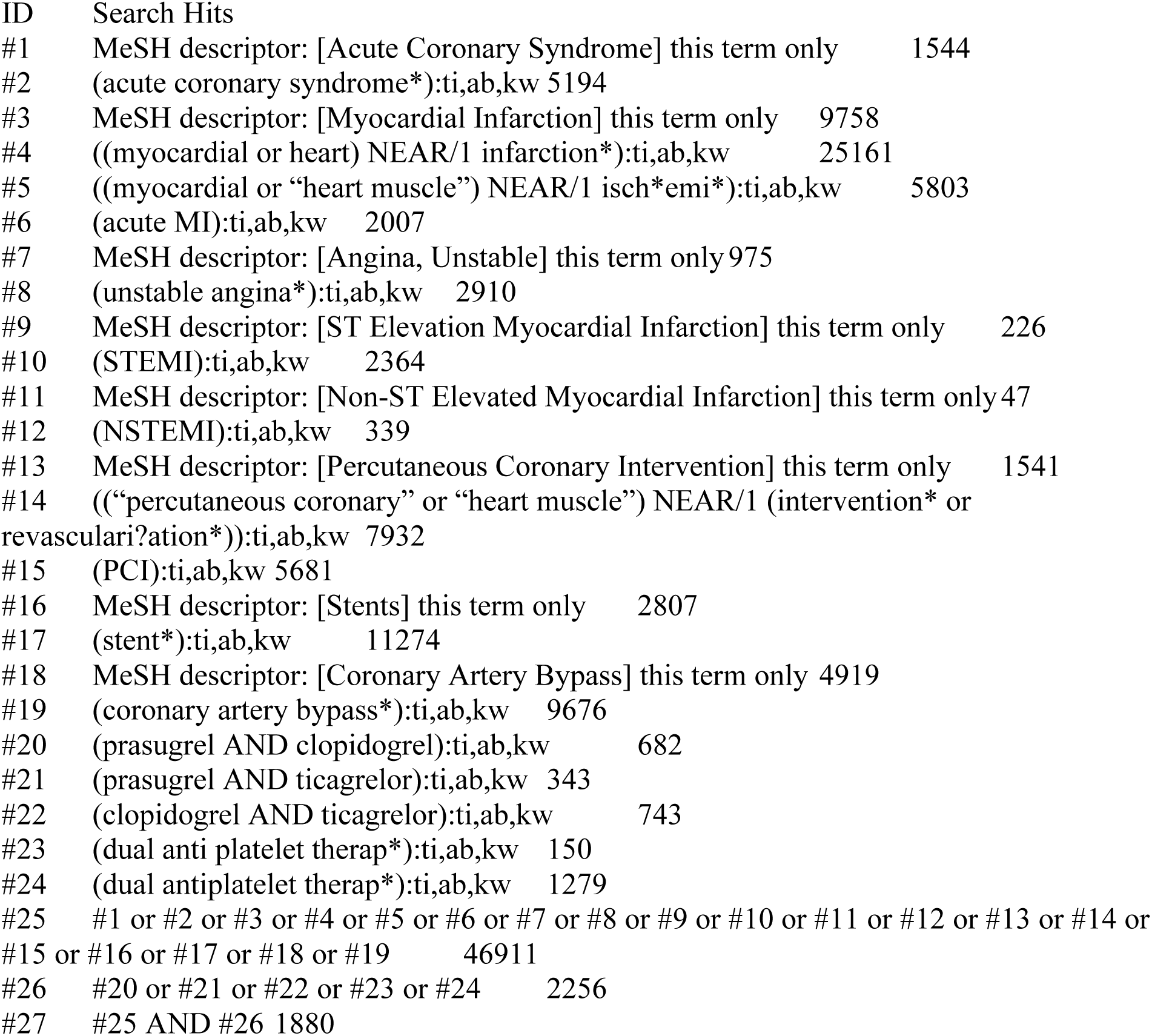

#### CLINICALTRIALS.GOV

##### Conditions or disease

((Acute Coronary Syndrome*) OR (myocardial infarction*) OR (myocardial ischemia*) OR (unstable angina*) OR (stemi) OR (nstemi) OR (percutaneous coronary) OR (stent*) OR (coronary artery bypass*))

##### Other terms

((clopidogrel AND ticagrelor) OR (prasugrel AND ticagrelor) OR (clopidogrel AND ticagrelor) OR (dual antiplatelet*) OR (dual anti platelet*) OR (DAPT))

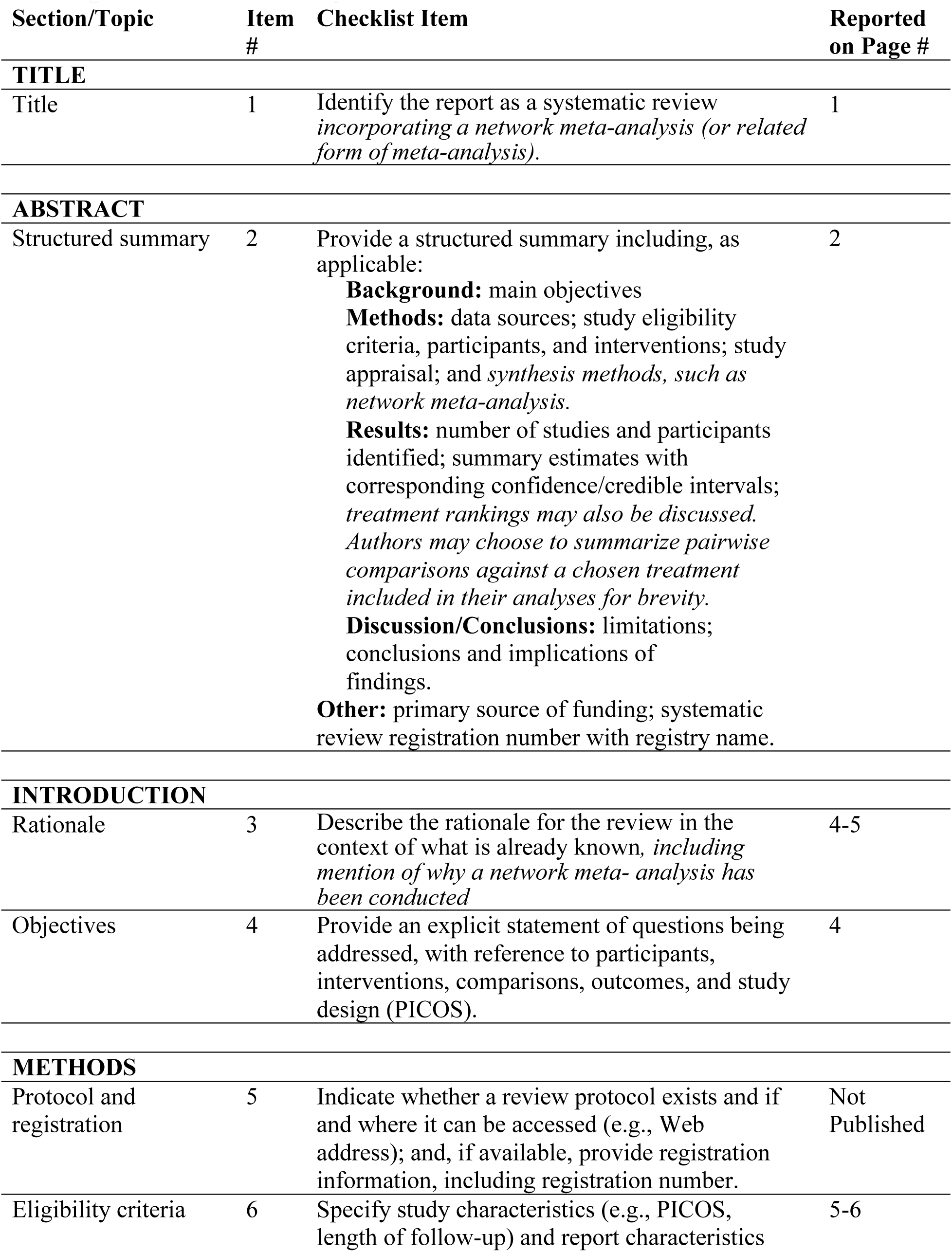
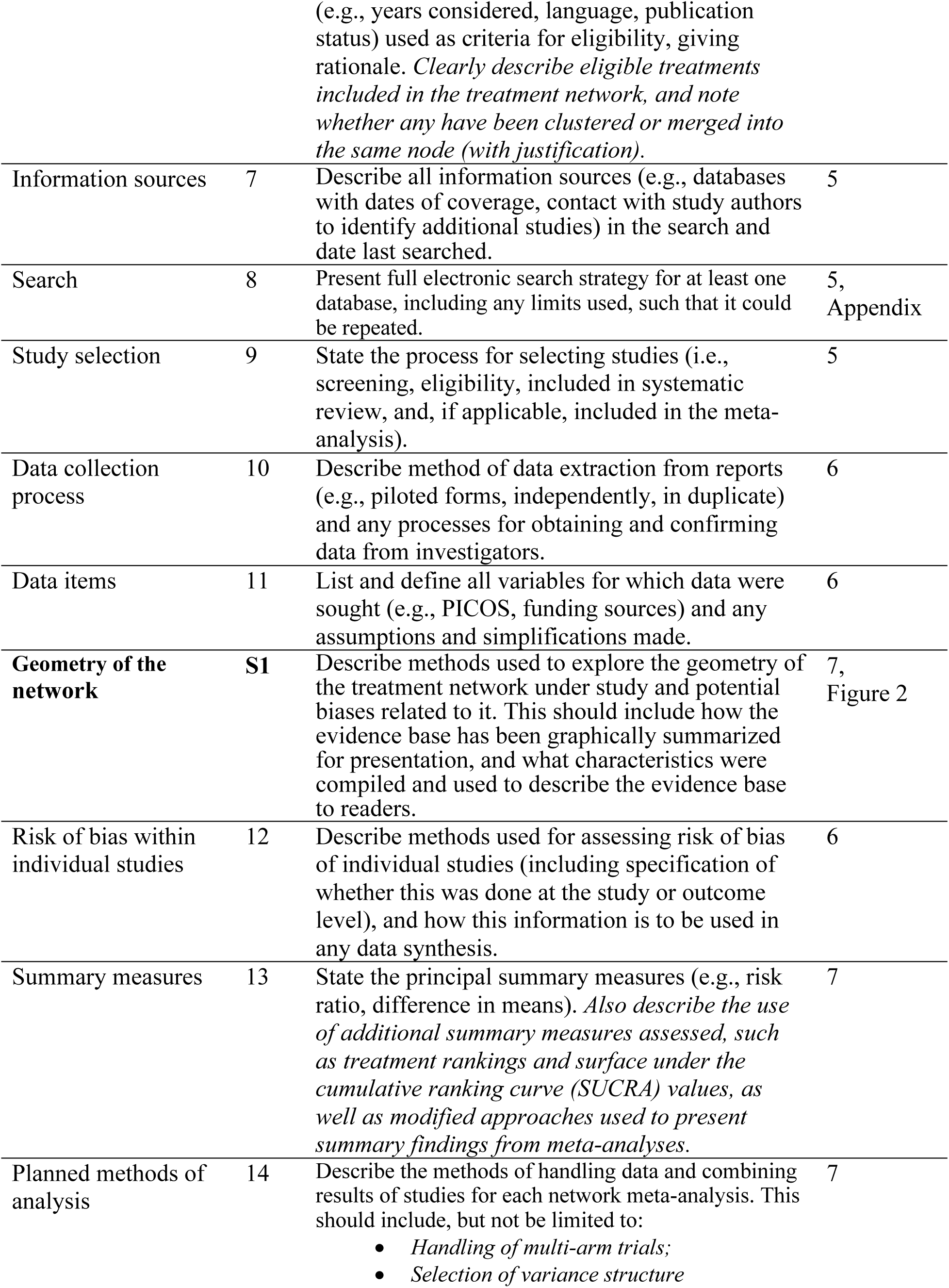
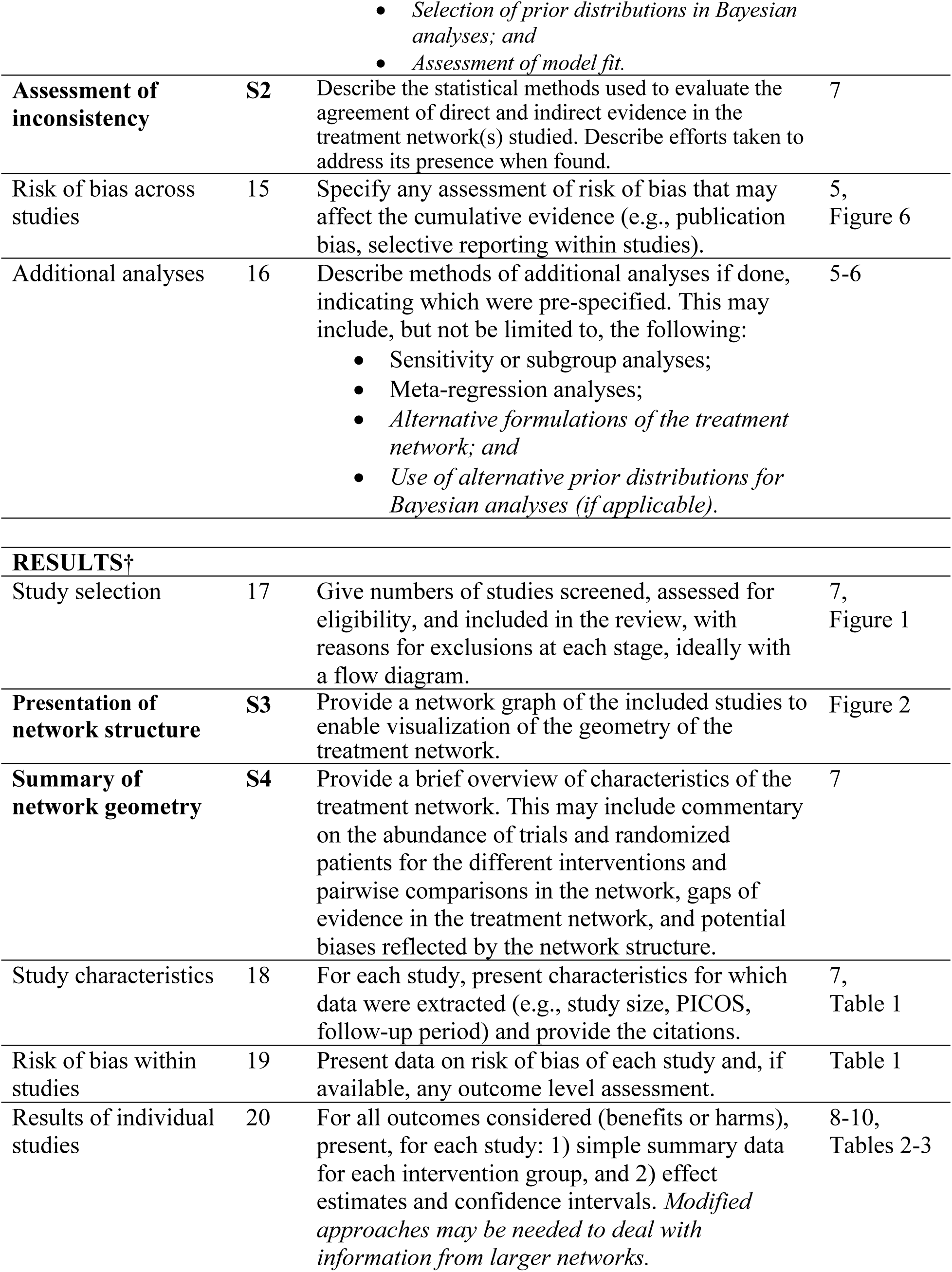
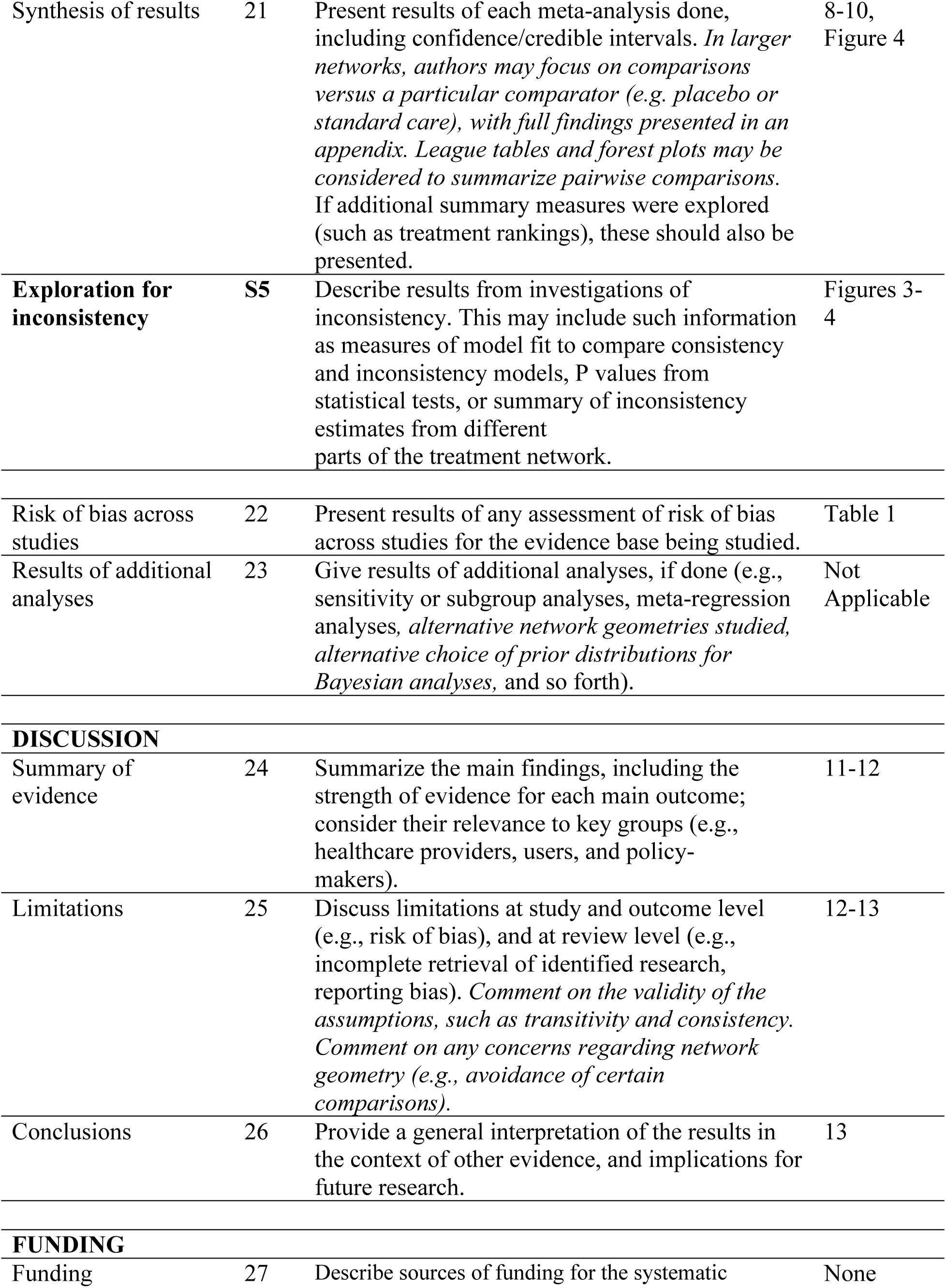
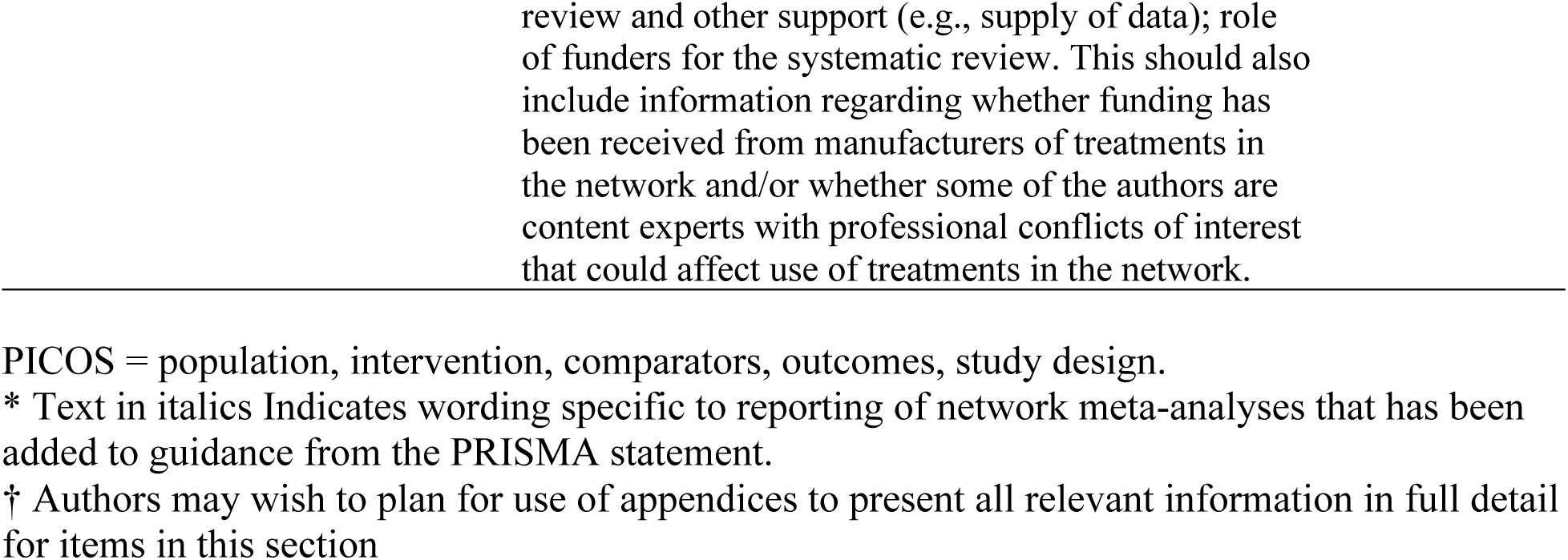
PRISMA NMA Checklist of Items to Include When Reporting A Systematic Review Involving a Network Meta-Analysis.

## REFERENCES

1. Levine GN, Bates ER, Bittl JA, et al. 2016 ACC/AHA Guideline Focused Update on Duration of Dual Antiplatelet Therapy in Patients With Coronary Artery Disease. J Am Coll Cardiol. 2016;68(10):1082–1115. doi:10.1016/j.jacc.2016.03.513

2. Mehta SR, Bainey KR, Cantor WJ, et al. 2018 Canadian Cardiovascular Society/Canadian Association of Interventional Cardiology Focused Update of the Guidelines for the Use of Antiplatelet Therapy. Can J Cardiol. 2018;34(3):214–233. doi:10.1016/j.cjca.2017.12.012

3. Bueno H, Byrne RA, Collet JP, et al. 2017 ESC focused update on dual antiplatelet therapy in coronary artery disease developed in collaboration with EACTS. :48.

4. Wallentin L, Becker RC, Budaj A, et al. Ticagrelor versus Clopidogrel in Patients with Acute Coronary Syndromes. N Engl J Med. 2009;361(11):1045–1057. doi:10.1056/NEJMoa0904327

5. Wiviott SD, Braunwald E, McCabe CH, et al. Prasugrel versus Clopidogrel in Patients with Acute Coronary Syndromes. N Engl J Med. 2007;357(20):2001–2015. doi:10.1056/NEJMoa0706482

6. Mahaffey KW, Wojdyla DM, Carroll K, et al. Ticagrelor Compared With Clopidogrel by Geographic Region in the Platelet Inhibition and Patient Outcomes (PLATO) Trial. Circulation. 2011;124(5):544–554. doi:10.1161/CIRCULATIONAHA.111.047498

7. Center for Drug Evaluation and Research, Complete Response Review Addendum Sponsor Safety Reporting Submissions: NDA 22-433 and IND 65,808 SD 632 Drug: Ticagrelor (BrilintaTM), Https://Www.Accessdata.Fda.Gov/Drugsatfda_docs/Nda/2011/022433orig1s000medr.Pdf.

8. Brophy JM. Multicenter trials, guidelines, and uncertainties — Do we know as much as we think we do? Int J Cardiol. 2015;187:600–603. doi:10.1016/j.ijcard.2015.04.004

9. Brophy JM. Key issues in the statistical interpretation of randomized clinical trials. Can J Cardiol. Published online January 2021:S0828282X20311806. doi:10.1016/j.cjca.2020.12.014

10. Hutton B, Salanti G, Caldwell DM, et al. The PRISMA Extension Statement for Reporting of Systematic Reviews Incorporating Network Meta-analyses of Health Care Interventions: Checklist and Explanations. Ann Intern Med. 2015;162(11):777–784. doi:10.7326/M14-2385

11. Sterne JAC, Savović J, Page MJ, et al. RoB 2: a revised tool for assessing risk of bias in randomised trials. Br Med J. 2019;366:I4898.

12. Dias S, Welton NJ, Caldwell DM, Ades AE. Checking consistency in mixed treatment comparison meta-analysis. Stat Med. 2010;29(7-8):932–944. doi:10.1002/sim.3767

13. Dias S, Welton NJ, Sutton AJ, Caldwell DM, Lu G, Ades AE. Evidence Synthesis for Decision Making 4: Inconsistency in Networks of Evidence Based on Randomized Controlled Trials. Med Decis Making. 2013;33(5):641–656. doi:10.1177/0272989X12455847

14. Dias S, Sutton AJ, Ades AE, Welton NJ. Evidence Synthesis for Decision Making 2: A Generalized Linear Modeling Framework for Pairwise and Network Meta-analysis of Randomized Controlled Trials. Med Decis Making. 2013;33(5):607–617. doi:10.1177/0272989X12458724

15. Gert van Valkenhoef and Joel Kuiper (2021). gemtc: Network Meta-Analysis Using Bayesian Methods. R package version 1.0–1. https://CRAN.R-project.org/package=gemtc.

16. Martyn Plummer (2022). rjags: Bayesian Graphical Models using MCMC. R package version 4–13. https://CRAN.R-project.org/package=rjags.

17. Brooks SP, Gelman A. General Methods for Monitoring Convergence of Iterative Simulations. J Comput Graph Stat. 1998;7(4):434–455. doi:10.1080/10618600.1998.10474787

18. Gelman A, Carlin JB, Stern HS, Dunson DB, Vehtari A, Rubin DB. Bayesian Data Analysis Third edition (with errors fixed as of 15 February 2021).

19. R Core Team (2020). R: A language and environment for statistical computing. R Foundation for Statistical Computing, Vienna, Austria. URL https://www.R-project.org/.

20. Wijeysundera DN, Austin PC, Hux JE, Beattie WS, Laupacis A. Bayesian statistical inference enhances the interpretation of contemporary randomized controlled trials. J Clin Epidemiol. 2009;62(1):13–21.e5. doi:10.1016/j.jclinepi.2008.07.006

21. Kruschke JK. Bayesian Analysis Reporting Guidelines. Nat Hum Behav. 2021;5(10):1282–1291. doi:10.1038/s41562-021-01177-7

22. Berwanger O, Lopes RD, Moia DDF, et al. Ticagrelor Versus Clopidogrel in Patients With STEMI Treated With Fibrinolysis. J Am Coll Cardiol. 2019;73(22):2819–2828. doi:10.1016/j.jacc.2019.03.011

23. Gasecka A, Nieuwland R, Budnik M, et al. Ticagrelor attenuates the increase of extracellular vesicle concentrations in plasma after acute myocardial infarction compared to clopidogrel. J Thromb Haemost JTH. 2020;18(3):609–623. doi:10.1111/jth.14689

24. Gimbel M, Qaderdan K, Willemsen L, et al. Clopidogrel versus ticagrelor or prasugrel in patients aged 70 years or older with non-ST-elevation acute coronary syndrome (POPular AGE): the randomised, open-label, non-inferiority trial. The Lancet. 2020;395(10233):1374–1381. doi:10.1016/S0140-6736%2820%2930325-1

25. Goto S, Huang CH, Park SJ, Emanuelsson H, Kimura T. Ticagrelor vs. Clopidogrel in Japanese, Korean and Taiwanese Patients With Acute Coronary Syndrome – Randomized, Double-Blind, Phase III PHILO Study –. Circ J. 2015;79(11):2452–2460. doi:10.1253/circj.CJ-15-0112

26. He P, Luo X, Li J, et al. Clinical Outcome between Ticagrelor versus Clopidogrel in Patients with Acute Coronary Syndrome and Diabetes. Puthanveetil P, ed. Cardiovasc Ther. 2021;2021:1–9. doi:10.1155/2021/5546260

27. Mohareb MW, AbdElghany M, Zaki HF, El-Abhar HS. Diabetes and CYP2C19 Polymorphism Synergistically Impair the Antiplatelet Activity of Clopidogrel Compared With Ticagrelor in Percutaneous Coronary Intervention-treated Acute Coronary Syndrome Patients. J Cardiovasc Pharmacol. 2020;76(4):478–488. doi:10.1097/FJC.0000000000000881

28. Park DW, Kwon O, Jang JS, et al. Clinically Significant Bleeding With Ticagrelor Versus Clopidogrel in Korean Patients With Acute Coronary Syndromes Intended for Invasive Management: A Randomized Clinical Trial. Circulation. 2019;140(23):1865–1877. doi:10.1161/CIRCULATIONAHA.119.041766

29. Wu X, You W, Wu Z, et al. Ticagrelor versus clopidogrel for prevention of subclinical stent thrombosis detected by optical coherence tomography in patients with drug-eluting stent implantation-a multicenter and randomized study. Platelets. 2021;32(3):404–412. doi:10.1080/09537104.2020.1754381

30. Roe M.T., Armstrong P.W., Fox K.A.A., et al. Prasugrel versus clopidogrel for acute coronary syndromes without revascularization. N Engl J Med. 2012;367(14):1297–1309. doi:10.1056/NEJMoa1205512

31. Saito S, Isshiki T, Kimura T, et al. Efficacy and Safety of Adjusted-Dose Prasugrel Compared With Clopidogrel in Japanese Patients With Acute Coronary Syndrome: – The PRASFIT-ACS Study –. Circ J. 2014;78(7):1684–1692. doi:10.1253/circj.CJ-13-1482

32. Savonitto S, Ferri LA, Piatti L, et al. Comparison of Reduced-Dose Prasugrel and Standard-Dose Clopidogrel in Elderly Patients With Acute Coronary Syndromes Undergoing Early Percutaneous Revascularization. Circulation. 2018;137(23):2435–2445. doi:10.1161/CIRCULATIONAHA.117.032180

33. Yabe T., Noike R., Okubo R., Amano H., Ikeda T. Infarct Size and Long-Term Clinical Outcomes of Prasugrel versus Clopidogrel in Patients with Acute Coronary Syndrome Undergoing Coronary Artery Stenting: A Prospective Randomized Study. Int J Angiol. 2022;((Yabe, Noike, Okubo, Amano, Ikeda) Department of Cardiovascular Medicine, Toho University, Faculty of Medicine, 6-11-1 Omorinishi, Ota-ku, Tokyo 143-8541, Japan). doi:10.1055/s-0042-1746417

34. Motovska Z, Hlinomaz O, Kala P, et al. 1-Year Outcomes of Patients Undergoing Primary Angioplasty for Myocardial Infarction Treated With Prasugrel Versus Ticagrelor. J Am Coll Cardiol. 2018;71(4):371–381. doi:10.1016/j.jacc.2017.11.008

35. Schupke S, Neumann FJ, Menichelli M, et al. Ticagrelor or Prasugrel in Patients with Acute Coronary Syndromes. N Engl J Med. 2019;381(16):1524–1534. doi:10.1056/NEJMoa1908973

36. van der Hoeven NW, Janssens GN, Everaars H, et al. Platelet Inhibition, Endothelial Function, and Clinical Outcome in Patients Presenting With ST-Segment-Elevation Myocardial Infarction Randomized to Ticagrelor Versus Prasugrel Maintenance Therapy: Long-Term Follow-Up of the REDUCE-MVI Trial. J Am Heart Assoc. 2020;9(5):e014411. doi:10.1161/JAHA.119.014411

37. Jin CD, Kim MH, Song K, et al. Pharmacodynamics and Outcomes of a De-Escalation Strategy with Half-Dose Prasugrel or Ticagrelor in East Asians Patients with Acute Coronary Syndrome: Results from HOPE-TAILOR Trial. J Clin Med. 2021;10(12). doi:10.3390/jcm10122699

38. Li DT, Li SB, Zheng JY, et al. Analysis of Ticagrelor’s Cardio-protective Effects on Patients with ST-segment Elevation Acute Coronary Syndrome Accompanied with Diabetes. Open Med Wars Pol. 2019;14(101672167):234–240. doi:10.1515/med-2019-0017

39. Lu Y, Li Y, Yao R, et al. Clinical effect of ticagrelor administered in acute coronary syndrome patients following percutaneous coronary intervention. Exp Ther Med. 2016;11(6):2177–2184.

40. Tang X, Li R, Jing Q, et al. Assessment of Ticagrelor Versus Clopidogrel Treatment in Patients With ST-elevation Myocardial Infarction Undergoing Primary Percutaneous Coronary Intervention. J Cardiovasc Pharmacol. 2016;68(2):115–120. doi:10.1097/FJC.0000000000000390

41. Wang H, Wang X. Efficacy and safety outcomes of ticagrelor compared with clopidogrel in Chinese elderly patients with acute coronary syndrome. Ther Clin Risk Manag. 2016;Volume 12:1101–1105. doi:10.2147/TCRM.S108965

42. Wang X, Li X, Wu H, et al. Beneficial effect of ticagrelor on microvascular perfusion in patients with ST-segment elevation myocardial infarction undergoing a primary percutaneous coronary intervention. Coron Artery Dis. 2019;30(5):317–322. doi:10.1097/MCA.0000000000000707

43. Wu HB, Tian HP, Wang XC, et al. Clinical efficacy of ticagrelor in patients undergoing emergency intervention for acute myocardial infarction and its impact on platelet aggregation rate. Am J Transl Res. 2018;10(7):2175–2183.

44. Yang B, Zheng C, Yu H, et al. Comparison of Ticagrelor and Clopidogrel for Patients Undergoing Emergency Percutaneous Coronary Intervention. Iran J Public Health. 2018;47(7):952–957.

45. Yao G, Su G, Li K, Li B, Dong H. Comparative study of ticagrelor and clopidogrel in therapeutic effect of acute myocardial infarction patients undergoing percutaneous coronary intervention. Saudi J Biol Sci. 2017;24(8):1818–1820. doi:10.1016/j.sjbs.2017.11.020

46. You J, Li H, Guo W, et al. Platelet function testing guided antiplatelet therapy reduces cardiovascular events in Chinese patients with ST-segment elevation myocardial infarction undergoing percutaneous coronary intervention: The PATROL study. Catheter Cardiovasc Interv. 2020;95(S1):598–605. doi:10.1002/ccd.28712

47. Zhang Y, Zhao Y, Pang M, et al. High-dose clopidogrel versus ticagrelor for treatment of acute coronary syndromes after percutaneous coronary intervention in CYP2C19 intermediate or poor metabolizers: a prospective, randomized, open-label, single-centre trial. Acta Cardiol. 2016;71(3):309–316. doi:10.1080/AC.71.3.3152091

48. Kitano D, Takayama T, Fukamachi D, et al. Impact of low-dose prasugrel on platelet reactivity and cardiac dysfunction in acute coronary syndrome patients requiring primary drug-eluting stent implantation: A randomized comparative study. Catheter Cardiovasc Interv Off J Soc Card Angiogr Interv. 2020;95(1):E8–E16. doi:10.1002/ccd.28277

49. Navarese EP, Khan SU, Kolodziejczak M, et al. Comparative Efficacy and Safety of Oral P2Y12 Inhibitors in Acute Coronary Syndrome: Network Meta-Analysis of 52 816 Patients From 12 Randomized Trials. Circulation. 2020;142(2):150–160. doi:10.1161/CIRCULATIONAHA.120.046786

50. Singh S, Singh M, Grewal N, Khosla S. Comparative Efficacy and Safety of Prasugrel, Ticagrelor, and Standard-Dose and High-Dose Clopidogrel in Patients Undergoing Percutaneous Coronary Intervention: A Network Meta-analysis. Am J Ther. Published online 2016.

51. Sterne JAC, Sutton AJ, Ioannidis JPA, et al. Recommendations for examining and interpreting funnel plot asymmetry in meta-analyses of randomised controlled trials. BMJ. 2011;343(jul22 1):d4002-d4002. doi:10.1136/bmj.d4002

